# Functional and microstructural brain abnormalities, fatigue, and cognitive dysfunction after mild COVID-19

**DOI:** 10.1101/2021.03.20.21253414

**Authors:** Lucas Scardua Silva, Rafael Batista Joao, Mateus Henrique Nogueira, Italo Karmann Aventurato, Brunno Machado de Campos, Mariana Rabelo de Brito, Marina Koutsodontis Machado Alvim, Guilherme Vieira Nunes Ludwig, Cristiane Rocha, Thierry Kaue Alves Silva Souza, Beatriz Amorim da Costa, Maria Julia Mendes, Takeshi Waku, Vinicius de Oliveira Boldrini, Natália Silva Brunetti, Sophia Nora Baptista, Gabriel da Silva Schmitt, Jhulia Gabriela Duarte de Sousa, Tânia Aparecida Marchiori de Oliveira Cardoso, André Schwambach Vieira, Leonilda Maria Barbosa Santos, Alessandro dos Santos Farias, Fernando Cendes, Clarissa Lin Yasuda

**Affiliations:** Brazilian Institute of Neuroscience and Neurotechnology (BRAINN), University of Campinas; Department of Neurology, Clinics Hospital, University of Campinas; Institute of Mathematics, Statistics and Scientific Computing, University of Campinas; Molecular Genetics Laboratory, Faculty of Medical Sciences, University of Campinas; Autoimmune Research Lab, Institute of Biology, University of Campinas; Department of Radiology, Clinics Hospital, University of Campinas

**Keywords:** COVID-19, neurocovid, SARS-CoV-2, neuroimaging, cognition, fatigue, resting-state

## Abstract

Although post-acute cognitive dysfunction and neuroimaging abnormalities have been reported after hospital discharge in patients recovered from COVID-19, little is known about persistent, long-term alterations in patients who did not require hospitalization. Therefore, we conducted a cross-sectional study of 87 consecutive, non-hospitalized recovered individuals, with a median of 54 days after the laboratory confirmation of COVID-19. We performed structured interviews, neurological examination, and 3T-MRI scans. The MRI study included white matter investigation with diffusion tensor images (DTI) and seed-based resting-state functional MRI (fMRI) connectivity analyses of the default mode network (DMN). In addition, we used validated instruments to examine fatigue, symptoms of anxiety and depression, somnolence, language, memory, and cognitive flexibility.

Individuals self-reported a high frequency of headaches (40%) and memory difficulties (33%). The quantitative analyses confirmed symptoms of fatigue (68% of participants), excessive somnolence (35%), symptoms of anxiety (29%), impaired cognitive flexibility (40%) and language dysfunction (33%). In addition, we observed a correlation between DTI fractional anisotropy (FA) and abnormal attention and cognitive flexibility measured by the Trail Making Test part B. The resting-state fMRI study of the DMN showed an association between higher connectivity of the posterior cingulate cortex (PCC) and higher levels of fatigue and somnolence. While greater connectivity of the PCC with bilateral angular gyri was associated with higher fatigue levels, the elevated levels of somnolence correlated with higher connectivity between the PCC and both the left thalamus and putamen.

## INTRODUCTION

Studies have consistently reported neurological manifestations of COVID-19. However, little is known about the long-term neurological events associated with the novel, severe acute respiratory syndrome coronavirus 2 (SARS-CoV-2)^1^. While most individuals will recover from respiratory symptoms, the long-term course of post-COVID-19 fatigue and cognitive dysfunction is uncertain. One French study identified dysexecutive syndrome in 15/45 (33%) patients with severe COVID-19 infection^2^.

Another Chinese study recruited 29 patients (after hospitalisation) and reported cognitive dysfunction after their recovery^3^. However, there is a limitation in the present understanding of the residual long-term neurological and cognitive dysfunctions (including the nature, duration, and pathophysiology) in individuals who recovered from COVID-19^4^, especially those who had a mild infection and who did not require hospitalisation.

Although the neuroinvasion of SARS-CoV-2 has been demonstrated with confirmation of the virus in brain autopsies^5^, the neural mechanisms underlying both neurological and neuropsychiatric symptoms (acute and chronic) remain unclear. One study with 60 patients (3 months after hospitalisation) identified grey and white matter abnormalities with MRI analyses^6^ but did not include cognitive tests. Given the lack of information about the long-term effects of COVID-19 after mild infection^4^, we investigated the nature of persistent neurological symptoms in this subgroup of non-hospitalised COVID-19 patients, combining clinical data with cognitive tests and structural and functional MRI analyses.

## METHODS

### Sample and study design

The Research Ethics Committee of the University of Campinas approved this study (Certificate of Ethical Appreciation Presentation - CAAE 31556920.0.0000.5404), and all subjects signed a consent form to participate.

#### Patients

We conducted a cross-sectional analysis of data from a longitudinal observational study designed to evaluate post-acute neurological symptoms and neuroimaging alterations related to COVID-19. We used social media to advertise our study with an online questionnaire^7^ (Supplementary Material, Table 14). We successively recruited the first 87 responders (who did not require hospitalisation and presented a confirmed diagnosis of COVID-19, Supplementary Figure 1) to visit our centre and to perform the four steps of the complete protocol:

a personal structured interview and neurological examination (performed by certified neurologists) and then

3T MRI acquisition, neuropsychological testing, and blood sample collection at the University of Campinas Hospital.

The COVID-19 diagnoses were based on polymerase chain reaction (PCR) tests or confirmed IgM or IgG antibodies.

As a control group, we recruited 55 healthy volunteers who did not present COVID-19 symptoms and had never tested positive for COVID-19 (median age 32 years [range 25-63]) from the same environment as the patients.

#### Neuropsychological evaluation

Due to the uncertainties related to cognitive impairment associated with SARS-CoV-2, we performed an exploratory neuropsychological evaluation of recovered individuals. We intentionally selected tests to evaluate different cognitive domains including *language* (the Verbal Categorical Fluency Test ^8^ and the Phonemic Verbal Fluency Test^9^), *episodic memory* (the Logical Memory subtest from the Wechsler Memory Scale [WMS-R]^10^) and *cognitive flexibility* (parts A and B of the Trail Making Test [TMT]^11^). We have included a detailed description in the Supplementary Material (the ‘Neuropsychological Instruments’ section).

We calculated the z-scores for the results of the neuropsychological tests based on the Brazilian standard and scaled scores. We controlled for the effects of age or schooling in a separate analysis using multiple linear regression residuals when normative data covered only one of these variables. For each test, the function was categorised as *preserved* (z-score > -0.66, including average, high-average, above-average, and exceptionally high scores), *low-average* score (z-score between -0.7 and - 1.26), *below-average* score (z-score between -1.32 and -1.82), and *exceptionally low* score (z-score < -1.96)^12^.

We quantified anxiety symptoms with the Beck Anxiety Inventory (BAI), and symptoms of depression with the Beck Depression Inventory II (BDI-II). We also investigated fatigue with the Chalder Fatigue Questionnaire (CFQ-11)^13 14^ and excessive daytime sleepiness with the Epworth Sleepiness Scale (ESS)^15^. Details of these tests are described in the Supplementary Material, the ‘Neuropsychological Instruments’ section.

#### MRI acquisitions

All individuals underwent 3T MRI (Phillips Achieva) with a 32-channel head coil. We acquired volumetric (3D) T1-weighted scans, resting-state functional MRI (fMRI) scans using echo-planar imaging (EPI) sequences, and diffusion tensor imaging (DTI) scans with 32 directions. The specific protocol for MRI acquisitions of COVID-19 individuals is included in the ‘MRI Protocol’ section of the Supplementary File.

#### MRI analyses

##### DTI analysis

We used a semi-automated tractography method based on a deterministic approach implemented in ExploreDTI (http://www.exploredti.com) and previously described in detail^16^. As an exploratory study, we used tractography to delineate different tracts, including the *commissural tracts* (corpus callosum divided into three parts: genu, body [BCC] and splenium [SPL]), *association tracts* (inferior longitudinal fasciculus [ILF], inferior fronto-occipital fasciculus [IFO] and uncinate fasciculus [UNC]), *limbic tracts* (dorsal cingulum [Dorsal-CING], parahippocampal cingulum [Parahippocampal-CING], and body of the fornix) and one *projection tract* (corticospinal tract [CST]). We averaged the diffusion parameters (fractional anisotropy [FA], mean diffusivity [MD], radial diffusivity [RD], and axial diffusivity [AD]) for each subject tract across all voxels reached by the tracts streamlines, ensuring that each voxel was counted only once ^16^. We included only tracts with at least five anatomically consistent streamlines^17^. We obtained right and left values separately for bilateral tracts, and we calculated a single value for each segment for the midline tracts (subdivisions of corpus callosum and fornix).

##### Functional connectivity (FC)

Given the uncertainty about the changes in cerebral functional FC after recovery from COVID-19 infection, we focussed the analyses on a well-known network, the default mode network (DMN)^18^. We investigated changes in the typical pattern observed in healthy volunteers, as well as the relationship between the DMN and fatigue^19^ and daytime sleepiness.

We performed the pre-processing steps and FC analysis with the UF^2^C toolbox (https://www.lniunicamp.com/uf2c) within SPM12 (http://www.fil.ion.ucl.ac.uk/spm/) running on MATLAB 2019b. For this analysis, we initially included 132 subjects: (55 controls (35 women, median age 32 years [range 25-63]) and 77 patients (59 women, median age 37 years [21-65]), paired for age (p=0.12) and sex (p=0.23). We have included detailed information about quality control and pre-processing in the ‘Functional Connectivity’ section of the Supplementary Material. The final number of subjects included in this analysis was 128: 52 controls (34 women, median age of 30 years, range 25-63) and 76 patients (59 women, median age of 37, range 21-65).

To investigate changes in the normal connectivity pattern of the DMN, we performed a study with the posterior cingulate cortex (PCC, 0 -51 21) as the seed. The time-series extracted from the region of interest (ROI) followed homogenisation procedures, excluding non-functionally representative voxels. The UF^2^C standard procedure excluded voxels with time-series that presented a low-outlier correlation with the seed-averaged time-series. We estimated the individual connectivity maps by using the Pearson correlation coefficient between the time-series from the PCC seed and all times-series of grey matter voxels. We converted these maps to z-score maps by using with Fisher’s r-score to z-score transformation. We used the generalised linear model (GLM) from SPM12 to perform second-level analyses for group inferences. We compared comparisons between patients and controls with a two-sample t-test controlling for sex and age. We also conducted separate linear regressions with individual maps and scores of fatigue (CFQ-11) and sleepiness (ESS), using sex and age as covariates.

The results were corrected for multiple comparisons. We initially applied a statistical threshold of p<0.001 (uncorrected at the voxel level, corresponding to T>3.16), with subsequent extent threshold at cluster level with Benjamini-Hochberg procedure^20^ (false discovery rate [FDR] corrected at p<0.05) to focus at the cluster level.

### Statistical analyses

We analysed the clinical data with SPSS 22. We used the chi-square test and Fisher’s exact test for categorical data. For continuous variables, we used the non-parametric Kruskal-Wallis and Mann-Whitney tests.

To compare the DTI data between patients and controls, we used separate models for each diffusion parameter (FA, MD, RD, and AD) covaried for age and sex, considering multivariate analysis of variance (MANOVA) for segments of the corpus callosum and the body of the fornix. For the bilateral tracts we applied repeated measures ANOVA (RM-ANOVA). We applied the Benjamini-Hochberg FDR adjustments^20^ adjustments with R^21^ to correct for multiple comparisons in each model^22^.

We calculated Pearson correlation coefficients between the neuropsychological data and DTI parameters for specific tracts according to the *a priori* hypothesis for each test, eliminating the need to correct for multiple comparisons. For language, we investigated the correlation between phonological fluency and FA of the ILF^23^. For the TMT, we investigated the association between the scores and the FA of the right ILF, as previously investigated in healthy-ageing individuals^24^. However, we were aware of some dissociations of DTI parameters and cognitive impairment, for example, alterations of mean diffusivity without abnormal FA^25^. Therefore, we explored the correlations between DTI indices (all four measurements) of both parahippocampal cingula (right and left sides) and episodic memory.

## RESULTS

### Clinical characteristics

We examined 87 individuals (64 women, median age 36 [range 18-71]), with a median interval between diagnosis and personal interview of 54 days (range 16-120 days). During the acute phase, patients reported a median of four symptoms (range 0-10, with five asymptomatic individuals), while during the post-acute stage (reported during the structured interview), they reported a median of two symptoms (range 0-13, with 25 asymptomatic individuals [28.7%]). The most frequent post-acute symptoms were fatigue (43.7%), headache (40%), memory difficulties (40%), anosmia (31%), and somnolence (18%). We described details about the types and percentages of these symptoms in Supplementary Figures 2 and 3. Interestingly, fatigue was reported by 38 individuals (43.7%) and was mostly combined with other symptoms such as headache (25 subjects), memory difficulties (17 subjects), and somnolence (14 subjects).

In terms of post-COVID-19 neurological examination and interview, we identified abnormalities in 11 (12.6%) individuals (described in Supplementary Table 1). One radiologist (JGDS) visually inspected all the structural MRI sacns and identified one haemangioma in the temporo-occipital region (the subject confirmed they had it before the infection); therefore, we excluded this subject from the MRI post-processing studies. The radiological evaluation of the MRI scans was normal for 86 (98.9%) individuals.

In addition to the structured interview, 65 subjects answered the CFQ-11 with a median of 15 points (range 0-32), and the ESS, with a median of 9 points (range 0-21). Differently from the proportion of symptoms reported during the interview (fatigue in 43.7% and somnolence in 18%), the binary classification (presence or absence of symptoms) resulting from the scores showed symptoms of fatigue in 44 of 65 individuals (68%) and excessive daytime sleepiness in 23 of 65 individuals (35%). The Pearson correlation coefficient between the CFQ-11 and ESS was moderate (r=0.44, p<0.001). Besides, excessive daytime sleepiness was more frequent in individuals with fatigue (20/44, 45%) than in those without fatigue (3/21, 14%, p=0.025).

### Neuropsychological evaluation

We performed the neuropsychological evaluation in a subset of 78 individuals (59 women, with a median age of 36 years [range 18-70]). Nine individuals did not undergo the evaluation due to a lack of time; no other reasons were reported. Approximately 18% of subjects presented depression symptoms (BDI-II >13), and 29% showed anxiety symptoms (BAI > 10). We identified a correlation between the BDI-II and CFQ-11 scores (r=0.47, p<0.001, controlled for BAI scores).

In terms of cognitive performance, we identified abnormal performance (below low and exceptionally low scores) in 33% of patients for phonological fluency, 30% of patients for the TMT-A, and 40% of patients from the TMT-B. (Figure 1, Supplementary Table 2).

**Figure1.**
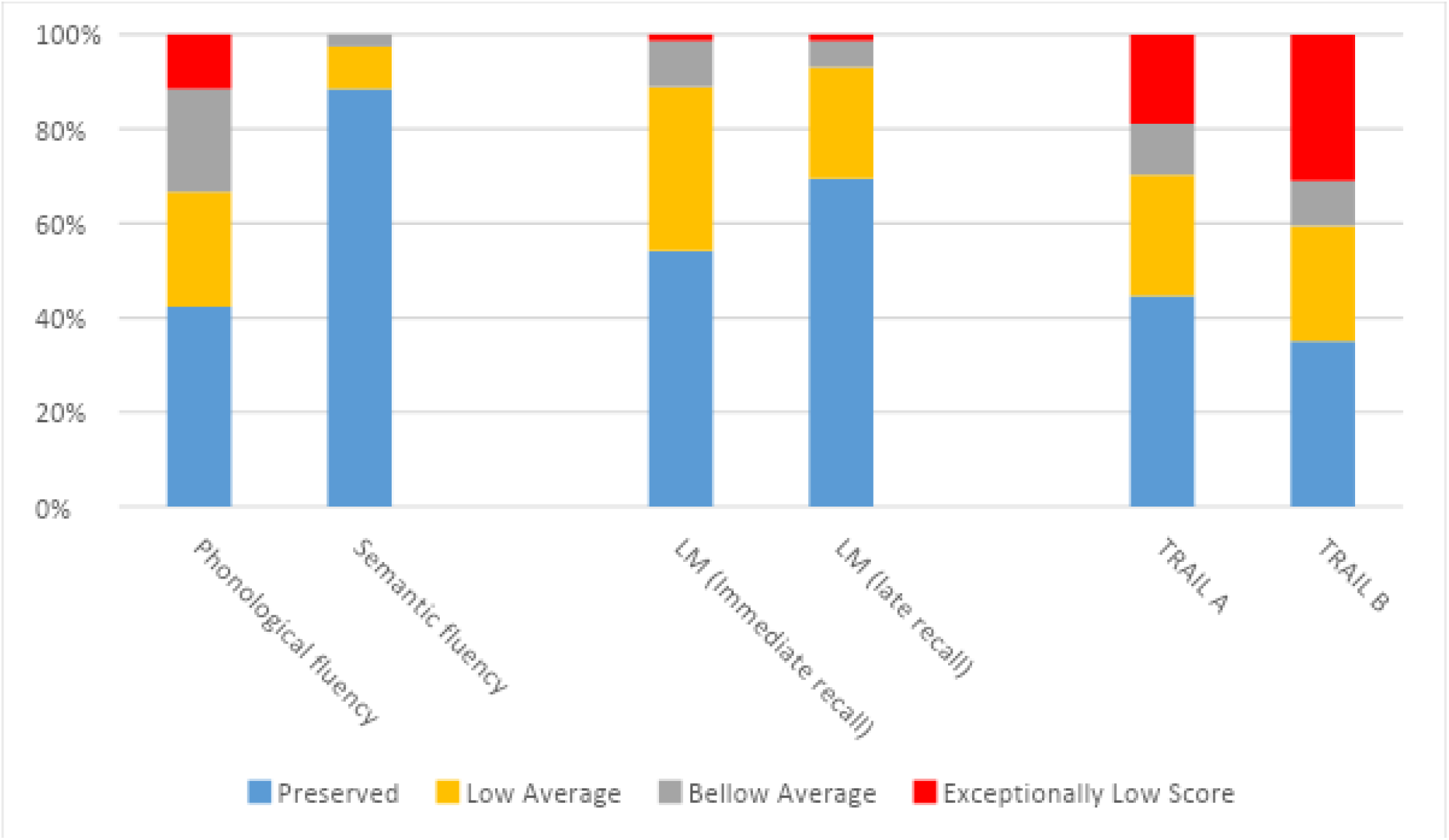
Neuropsychological evaluation of patients recovered from (mild) COVID-19 shows cognitive dysfunction mainly in parts A and B of the Trail Making Test and verbal fluency. LM, logical memory test.

### Neuroimaging findings

#### DTI results

We analysed 80 post-COVID patients and 55 controls paired for age (p=0.094) and sex (p=0.30). As an exploratory study, we identified a trend for higher FA values for patients, associated with lower MD, RD, and AD values (with some exceptions). However, the results did not survive the corrections for multiple comparisons. Full statistical details and results are presented in Supplementary Tables 3-10

#### Correlations between DTI parameters and neuropsychological scores

Because we had paired neuropsychological and DTI data from 71 participants, we investigated the relationship between diffusivities and neuropsychological performance based on previous studies. The FA of the right ILF was mildly correlated with the TMT-B (r=0.3, p=0.015). However, we did not identify correlations between the ILF and phonological fluency or neither between the logical memory scores (immediate and delayed recall) and diffusivities in the parahippocampal cinguli.

#### FC of the DMN

We compared 76 patients and 52 controls, paired for age (p=0.072) and sex (p=0.127).

Although differences between the maps of FC from patients and controls did not survive the FDR correction, there was a trend towards reduced connectivity in the group recovered from COVID-19 in the left insula and right occipital region. (Supplementary Table 11).

Given the high proportion of subjects with symptoms of fatigue and sleepiness and previous associations between these symptoms and the DMN^26^, we further investigated the relationship between the DMN maps and scores of fatigue (CFQ-11; median 16, range 0-32) ^13^ and sleepiness ^27^ (ESS, median 8, range 0-21) ^15^ in 59 individuals. We performed two separate linear regressions between the FC maps and the ESS and CFQ-11. There was a positive correlation between the CFQ-11 scores and increased FC between the PCC and the left angular gyrus (−46, -64, 36) and the right angular gyrus (42, -56, 22) (Figure 2A). There was also a positive correlation between the ESS scores and increased FC between the PCC and the left thalamus (−8, -20, 18) and the left putamen (−22, 10, 0) (Figure 2B and Supplementary Figures 12 and 13).

**Figure 2.**
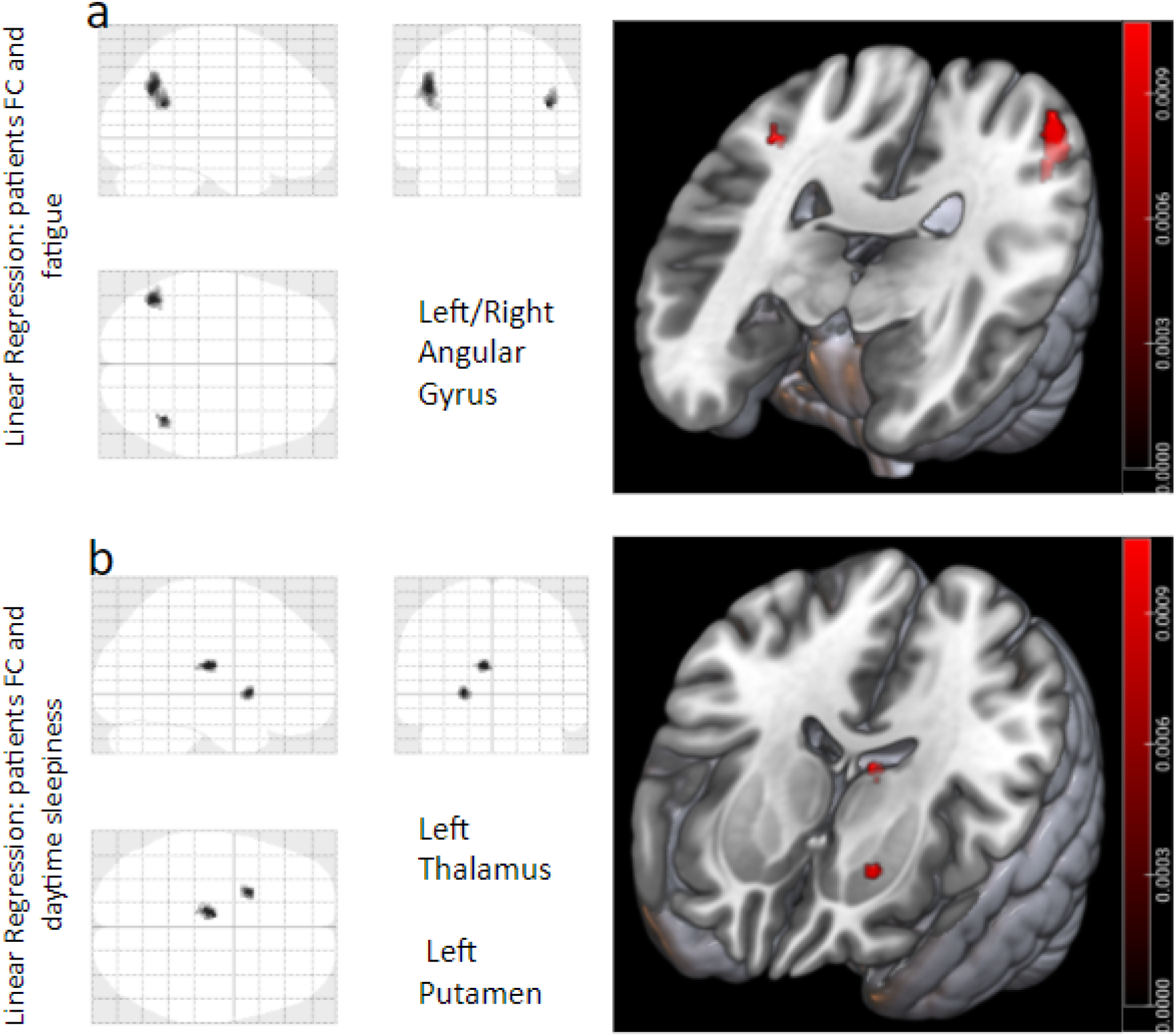
Seed-based functional MRI (fMRI) analyses of patients recovered from COVID-19; the seed is in the posterior cingulate cortex (0, -51, 21). The tests were covariated with sex and age. The structure names are based on the Neuromorphometrics Atlas. Only p<0.001, (after FDRc correction), was considered statistically significant. **a)** Linear regression between the patients’ functional connectivity (FC) maps and Chalder Fatigue Questionnaire scores revealed a positive correlation in the left angular gyrus (−46, -64, 36) and the right angular gyrus (42, -56, 22); **b)** Linear regression between FC maps and their Epworth Sleepiness Scale scores showed a positive correlation in the left thalamus (−8, -20, 18) and the left putamen (−22, 10, 0).

## DISCUSSION

We detected persistent headache, fatigue, excessive somnolence, anxiety, and cognitive dysfunction along with subtle structural and functional brain MRI abnormalities 2 months after the acute COVID-19 infection of non-hospitalized individuals. We identified a correlation between white matter alteration and abnormal attention and cognitive flexibility. In addition, there were alterations in brain connectivity of the DMN associated with excessive sleepiness and fatigue, indicating a complex pattern of altered cerebral functional networks.

### Persistent clinical symptoms

Two months after the acute period of COVID-19 infection, the most frequently reported symptoms were fatigue (43.7%), headache (40%), and memory difficulties (33%), similarly to the results from a preprint meta-analysis^28^. Interestingly, the CFQ-11 and ESS quantitative analysis revealed a higher proportion of symptomatic individuals (68% with fatigue and 35% with excessive daytime somnolence) compared with the proportions of symptoms reported during the structured interview.

Furthermore, while the presence of depression symptoms (18% of subjects, according to the BDI-II scores) was higher than what the patients self-reported (1%), the frequency of anxiety symptoms (29% of subjects) according to the BAI scores was close to those self-reported (23%). Overall, these findings are in accordance with a previous study^14^, as we observed a similar median score of the CFQ-11 (median of 16 points) in our group of non-hospitalised participants. While we detected a positive correlation between fatigue scores and the intensity of depression symptoms (BDI-II scores), we also described the elevated proportion of individuals with fatigue in participants with a history of anxiety/depression.

The proportion of female responders was higher than male responders, probably due to our recruitment method. There is still no clear evidence regarding whether sex is a risk factor for developing or perpetuating the long-term effects of COVID-19^28^; however, some studies have demonstrated that post-infection fatigue seems to be more frequent in women^14^. Hence, it is possible that we observed high rates of fatigue in our study because more affected women were interested in enrolling than men.

### Cognitive dysfunction and COVID-19

Recent studies have confirmed long-term cognitive dysfunction in survivors of COVID-19 (from ICU and ward hospitalisation)^3 29^. However, cognitive impairment after mild infection (without hospitalisation) is not well understood. In a recent analysis of 35 patients after hospital discharge (12.6 years of education and approximately 26 days after discharge), the application of supervised neuropsychological tests revealed abnormal performances mainly in verbal fluency (11.4%), mental flexibility and working memory (8.6%)^29^. Our group of 78 non-hospitalised participants presented higher rates of abnormal phonemic fluency (33%) and a high proportion of dysfunction in mental flexibility (40% showed impaired TMT-B test performance). Compared with the above-mentioned study, we examine more participants approximately 54 days after the acute period. Given the uncertainties related to the new virus, there is the possibility that cognitive dysfunction increases over time. So far, other researchers have applied a variety of non-supervised cognitive tests^3 30^ and also have demonstrated attention deficits in survivors. Despite the methodological differences, one study^30^ evaluated a large sample of individuals (although only 361 [0.42%] had a confirmed COVID-19 diagnosis) and detected abnormalities in ‘higher cognitive or executive function’, especially in tasks with a semantic component, and in those requiring selective visual attention.

It was surprising in our data that a high proportion of subjects (with a median education of 15 years) performed poorly, specifically in the TMT. This outcome can be partially explained by our convenience sample, which may have attracted more symptomatic participants. Besides, the poor performance on the TMT-B test correlated with abnormalities in the white matter structure (reduced FA) of the right ILF. A similar association has been reported in a study on healthy ageing^24^, which has been associated with visual processing^31^. It also correlated with a thinner cortex in the rectus gyrus (submitted data).

### MRI analyses

#### DTI alterations

While one recent study of six patients with severe disease identified overall reduced FA associated with increased MD values^32^, another study with 60 hospitalised patients observed higher FA associated with reduced MD, RD and AD^6^. Interestingly, we observed similar trends in our diffusivities, although our patients did not require hospitalisation. These findings suggest a negative impact of the virus on the nervous system regardless of the severity of the acute stage. While the reduced FA in the acute phase may suggest overall white matter damage, elevated FA in the post-acute stage may suggest the neuroplasticity phenomenon as observed in neurodegenerative disease^33^.

Due to the cognitive impairment confirmed with traditional tests, we were intrigued by the poor performance on the TMT-B. Therefore, we investigated possible neural correlates with the right ILF, as described by Perry et al^24^. We found an association between altered FA of the right (but not the left) ILF and the TRAIL B z-scores (adjusted for age, sex and education). Because the ILF is a long brain tract that connects the temporal and occipital lobes and has a role in visual cognition^34^, and visual memory^35^, we hypothesise that its deterioration may contribute to the difficulties in the set-shifting tests.

#### FC analyses

We identified increased FC between the PCC and angular gyri, that correlated with fatigue scores. This is an intriguing finding because it has been observed in a study of chronic fatigue syndrome (CFS)^19^ and in another with cognitive fatigue induced in 39 healthy individuals^36^. Besides, atrophy in the angular gyrus has been reported in a study with 26 patients with CFS^37^. However, considering the short time interval between the acute infection and MRI acquisition, it was surprising to detect abnormal FC in patients with a mild infection, because the fatigue intensity was variable and most patients did not fulfill the criteria for CFS^37^. Nevertheless, our data supports the hypothesis of neurological involvement in the physiopathology of fatigue.

We identified a relationship between sleepiness and alterations in the DMN, as has been demonstrated in idiopathic hypersomnia^26^ and sleep deprivation^38^. More specifically, we found a positive correlation between PCC connection with the thalamus and putamen and daytime sleepiness in patients recovered from COVID-19. Our results are consistent with recent evidence about the contribution of the thalamic connections to both sleepiness and disorders of consciousness in general ^38^. Moreover, recent studies have discussed the importance of the basal ganglia in sleepiness^39^ and wakefulness^40^.

Considering our results, a candidate theory that unifies the symptoms and network changes in post-COVID-19 patients must consider the DMN and its relations.

Unfortunately, we cannot explain the underlying pathophysiological mechanisms. We also cannot predict the duration of such states in the post-infected individuals. However, we are performing longitudinal analyses that will allow us to comprehend how transient or permanent these changes are and provide more evidence related to possible undesired and unexpected consequences of these persistent changes in brain dynamics networks.

## LIMITATIONS

The nature of this study - a cross-sectional design with a convenience sample - has restrained our ability to generalise of our findings. However, we believe our initial observations raise concerns about possible long-term impairment in patients recovered from mild COVID-19, especially considering the limited understanding of the neurological impact of SARS-CoV-2 infection. Nevertheless, our longitudinal analyses with a larger sample will provide additional insight regarding these long-term, persistent symptoms related to mood, cognition and brain connectivity.

## CONCLUSIONS

Our findings suggest the SARS-CoV-2 affects the brain in individuals who did not require hospitalisation, with persistent fatigue, headache, memory problems, and somnolence present even 2 months after their COVID-19 diagnosis. We detected cognitive impairment in these individuals, along with subtle white matter and connectivity abnormalities. The brain alterations and the severity of cognitive dysfunction increase need for extensive longitudinal studies of chronic neuropsychiatric symptoms in patients recovered from COVID-19, even in those with mild symptoms in the acute phase. Specific treatment of symptoms and neurorehabilitation strategies may be necessary to improve the quality of life and cognitive function for those with persistent limitations after the acute phase.

## Data Availability

Data collected and analyzed for this study will be available at the University database. Neuroimaging raw data will be available upon reasonable request.

## Data availability

Data collected and analysed for this study will be available in the University of Campinas database. The raw neuroimaging data will be available upon reasonable request.

## Acknowledgments

We thank the team of assistants who worked on the consecutive recruitment of patients: Andrea Ismara de Araújo Ruas, Lilian Cristina dos Santos Capelli and Sonia Neves Romeu Silva. We are also grateful for the work of the radiology team.

