## supplemental for "Functional and microstructural brain abnormalities, fatigue, and cognitive dysfunction after mild COVID-19"

Supplementary File

**Material and Methods**

**Supplementary Figure 1.** Flowchart with steps following the recruited participants

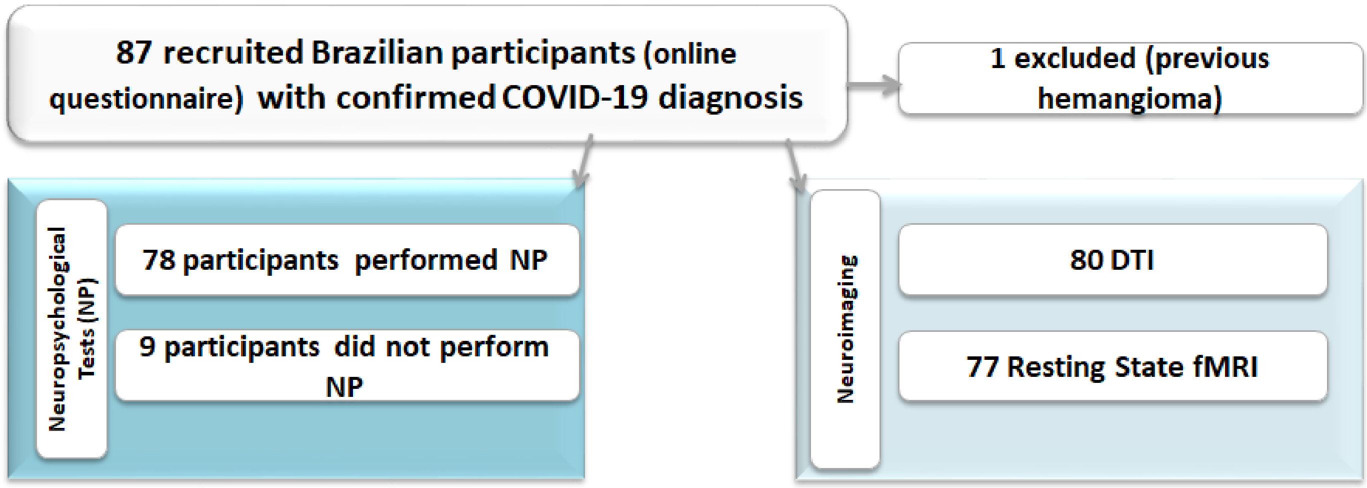

<u>Neuropsychological Instruments</u>

The **Verbal Categorical Fluency Test** [1] [2] is a verbal fluency task encompassing a semantic category (animals). This test was introduced as part of the Boston Diagnostic Aphasia Examination and can be used separately to evaluate the language, semantic memory, and cognitive flexibility. This task consists of recalling the highest number of animals in 60 seconds, and its score refers to the total amount of words about the topic (animals).

The **Phonemic Verbal Fluency Test**[3] consists of a naming words task that aims at recollecting words beginning with the letters F, A, and S, respectively, with a one-minute threshold for each letter. The final score is the sum of the words spoken correctly in each trial. This task evaluates language, semantic memory, and executive function.

The **Logical Memory subtest** from the Wechsler Memory Scale (WMS-R)[4 5] is a worldwide test used to evaluate immediate and delayed episodic memory. The examiner verbally presents two stories, and each story includes 25 pertinent pieces of information. Subjects are required to recall details of each story immediately after its presentation and again after 20 minutes.

The **Trail Making Test** (TMT)[6 7] is subdivided into two parts. Part A assesses processing speed and visual search in a task that requires ascending connection order of 25 numbers, randomly arranged. Part B evaluates alternating attention and cognitive flexibility in a task associated with shifting rules in an ascending sequence of 25 numbers. A training stage is applied to both parts.

The **Beck Depression Inventory (BDI-II)**[8] is a self-administered scale used for screening the severity of depressive symptoms. It consists of a standard multiple-choice questionnaire with 21 questions containing four items each. The total score ranges from 0 to 63 and the levels of severity thresholds follow 0-13 for minimal or no depression; 14-19 for mild depression; 20-28 for moderate depression; and 29-63 for severe depression symptoms.

The **Beck Anxiety Inventory (BAI)**[9] is also a self-administered scale for screening the severity of anxiety symptoms. This scale consists of 21 items with descriptive statements of physical and subjective symptoms of anxiety. The total score ranges from 0 to 63, allowing the characterization of intensity levels according to the following cutoffs: 0-10 for minimal or asymptomatic anxiety symptoms; 11-19 as mild anxiety; 20-30 as moderate anxiety; and 31-63 as severe anxiety symptoms.

The **Chalder Fatigue Scale** (CFQ-11)[10] questionnaire was used to assess the severity of fatigue. The participants answer 11 items on a 4-point scale (0-3). The global score range from 0-33. The scale can be used to separate “cases” and “non-cases”, based on a binary fatigue score ranging from 0-11. According to the binary system, scores of 4 or more are considered “cases” with fatigue[11].

We used the **Epworth sleepiness scale (ESS)** to estimate excessive daytime sleepiness (EDS). It is a self-report questionnaire with 8 situations involving daily activities. The final global scores range from 0-24; the diagnosis of EDS is suggested for those with scores higher than 10[12].

MRI Protocol (3T Philips-Achieva) designed for Post-COVID individuals:

- Axial diffusion weighted image (DWI) with acquisition voxel sizes of 1.51x1.95x3 $mm^3$ reconstructed with 0.9x0.9x3 $mm^3$, 55 slices, gap = 0.3 mm, TR = 3776 ms, TE = 94 ms, flip angle = 90º and FOV = 230x230 $mm^2$.
- Axial susceptibility weighted image (SWI) with acquisition voxel sizes of 0.6x0.6x2 $mm^3$ reconstructed with 0.34x0.34x1 $mm^3$, 142 slices, no gap, TR = 42 ms, 6 echos, first echo = 7.19 ms and echospacing = 6.2 ms, flip angle = 17º and FOV = 230x189 $mm^2$.
- Sagittal T1 3D WI with isotropic voxels of 1 mm, acquired in the sagittal plane, 1 mm thick, no gap, flip angle=8°, TR=7.0 ms, TE=3.2 ms, matrix=240x240, FOV=240x240 $mm^2$

- Sagittal T2-3D fluid attenuated inversion recovery image (FLAIR) with isotropic voxels of 1.2 mm, reconstructed with 0.5x0.5x0.6 $mm^3$, 300 slices, gap = 0.6 mm, TR=10000 ms, TI = 2680 ms, TE=276 ms, 2 averages (sampling averages), FOV = 250x250 mm2
- Resting-state: Echo planar images (EPI) with voxel sizes of 3x3x3 $mm^3$, acquired on the axial plane with 40 slices, no gap, flip angle=90º, TR=2 s, TE=30 ms in a 6-minute scan resulting in 180 dynamics and FOV = 240x240 $mm^2$.
- DWI/DTI: Diffusion tensor image (multiple diffusion direction images) with acquisition voxel sizes of 2x2x2 $mm^3$ reconstructed with 1x1x2 $mm^3$, 70 slices,

no gap, TR = 8500 ms, TE = 61 ms, flip angle 90°; 32 gradient directions; no averages; max b-factor = 1000 s/mm², FOV = 256x256 mm².

Functional connectivity

We performed the functional connectivity study using the UF²C toolbox (https://www.lniunicamp.com/uf2c) within SPM12 (http://www.fil.ion.ucl.ac.uk/spm/, using MATLAB 2019b) [13]. For this analysis, we initially included 132 subjects (55 controls and 77 patients). From the original group of 87, one individual was excluded due to previous large hemangioma and three due to movements' artifacts. The images preprocessing followed the UF²C standard pipeline and briefly, was based on functional image realignment, normalization to the MNI-space (Montreal Neurologic Institute Standard template), co-registration with T1-WI image, framewise displacement (FD), and the derivative variance (DVAR) estimation (head movement parameters) and smoothing with a kernel of 6x6x6 mm (FWHM). Additionally, structural images segmentation, modulation, and normalization (MNI space). After the initial preprocessing, the functional images were detrended (to remove MR linear trend), band-pass filtered (0.008-0.1 Hz), and regressed for censoring vectors (framewise displacement (FD) >0.5mm), white matter (WM), and CSF average signal, and for six realignment parameters.

- We performed the following quality control procedures:

Using the estimated individual FD, we excluded three subjects (2 controls and 1 patient) who presented abnormal values for FD (severe upper outliers [values higher than one interquartile range (IQR) regarding the volunteers mean value]). One control was excluded due to an image error. After this procedure, the average FD for the control group was 0.180 mm and 0.175 mm for the patients' group. No statistical difference was found between groups (FD, p = 0.716, Mann-Whitney test). Aiming to guarantee the intersubject FOV reproducibility, we used the $UF^2C$ outlier tool with the post-processed functional images: individual binary masks were created with the post-processed functional mean image of each volunteer, then, the tool identified any subjects with an abnormal (higher than 1.5*IQR) number of voxels that were outside the group averaged mask. The final number of subjects included in the analysis was 128 [52 controls (34 women, median age of 30 years, range 25-63) and 76 patients (59 women, median age of 37, range 21-65)].

Supplementary results

Supplementary Figure 2. The graph shows the reported symptoms in the COVID-19 acute phase of 87 patients.

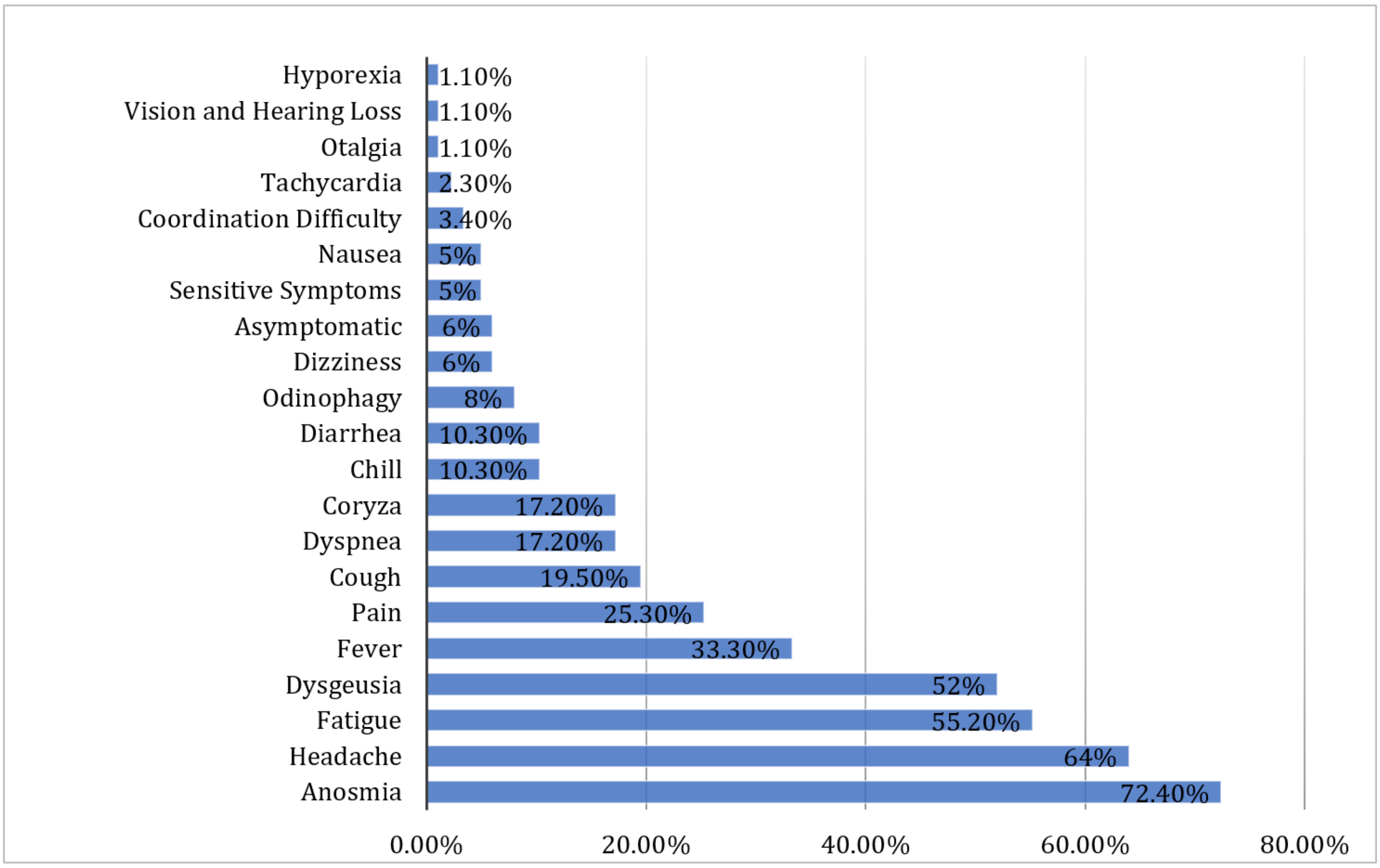

Supplementary Figure 3. The graph shows the reported symptoms of 87 patients approximately 54 days after the COVID-19 acute stage.

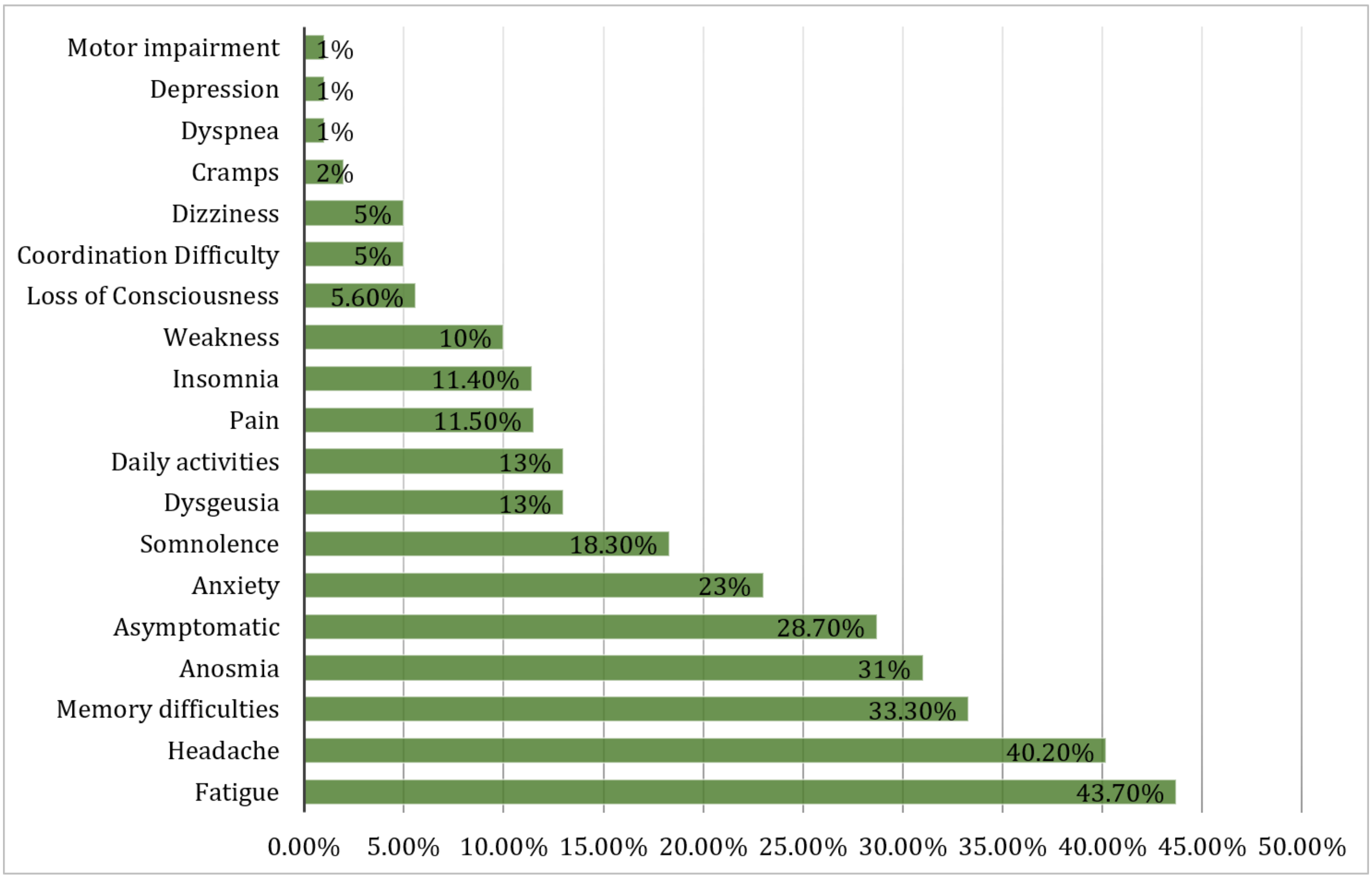

Supplementary Table 1. Description of participants with abnormalities in the neurological examination.

| Gender | Age (Decade of Life) | Neurological finding |
|---|---|---|
| Male | 40-49 | Spastic paraparesis; myalgia. |
| Male | 70-79 | Somnolence (Glasgow Coma Scale 14 (EOR4 VR 4 M 6)); poverty of speech; upper limb strength 3/5 and lower limb strength 2/5 |
| Female | 50-59 | Paresthesia in the lower left face; myalgia |
| Female | 40-49 | Dysdiadocokinesia |
| Female | 30-39 | Nystagmus; dizziness (benign paroxysmal positional vertigo (BPPV) like) |
| Male | 20-29 | Distal hypopalesthesia in the four limbs |
| Female | 50-59 | Hyperreflexia in the left superior limb |
| Male | 20-29 | Hypoesthesia of the right superior limb |
| Male | 50-59 | Paraparesis, cerebellar ataxia |
| Male | 50-59 | Intention and rest tremor; cerebellar ataxia |
| Female | 60-69 | Sensory ataxia; Tromner sign in the right hand; slowness of right hand |

| Supplementary Table 2. Neuropsychological evaluation of post-COVID 19 patients (78 individuals) | | |
|---|---|---|
| | **Median** | **Interval** |
| Age | 36 | [18–70] |
| Education (years) | 15 | [6–24] |
| BDI-II Score | 6.5 | [0–36] |
| BAI Score | 6 | [0–45] |
| CFQ-11 Score (59 subjects) | 16 | [0–32] |
| ESS (59 subjects) | 9 | [0–21] |
| **Neuropsychological tests** | | |
| | **Median of Z-score** | **Interval (min-max)** |
| **Phonological Fluency** | -0.98 | [-2.76 to 2.37] |

| | | |
|---|---|---|
| **Semantic Fluency** | 0.41 | [-1.52 to 2.73] |
| **Logical Memory (Immediate recall)** | -0.63 | [-2.26 to 1.96] |
| **Logical Memory (delayed recall)** | -0.3 | [-2.38 to 1.57] |
| **Trail Test** | | |
| A | -0.74 | [-7.05 to 1.38] |
| B | -1.1 | [-5.55 to 1.25] |
| BDI: Beck depression inventory; BAI: Beck anxiety inventory; CFQ: Chalder Fatigue Questionnaire; ESS: Epworth Sleepiness Scale; | | |

**DTI RESULTS**

**Fractional Anisotropy (FA)**

Supplementary Table 3. For FA, after a multivariate analysis of variance (MANOVA), we did not identify significant interaction between tracts × groups (Pillai's Trace = 0.022, $F_{[4,128]}$ =0.732, p=0.572, partial $\eta^2$ = 0.022, Observed Power = 0.231). Data presented in the table show the trend for higher FA values for the patients.

| **FA -Tract** | | **Mean** | **Std. Error** | **95% Confidence Interval** | |
|---|---|---|---|---|---|
| | | | | **Low. Bound** | **Upp. Bound** |
| **Body CC** | **Controls** | 0.574 | 0.002 | 0.570 | 0.578 |
| | **Patients** | 0.578 | 0.002 | 0.574 | 0.582 |
| **Fornix** | **Controls** | 0.453 | 0.003 | 0.447 | 0.459 |
| | **Patients** | 0.456 | 0.003 | 0.451 | 0.461 |
| **Genu CC** | **Controls** | 0.534 | 0.002 | 0.529 | 0.538 |
| | **Patients** | 0.538 | 0.002 | 0.534 | 0.542 |
| **Splenium CC** | **Controls** | 0.580 | 0.002 | 0.577 | 0.584 |
| | **Patients** | 0.583 | 0.002 | 0.580 | 0.586 |

*Comparison (MANOVA) of FA in midline tracts of post-COVID patients and controls. Besides no statistical difference, post-COVID patients present a trend of higher FA values in midline tracts.*
*FA: Fractional Anisotropy; CC: Corpus Callosum*

Supplementary Table 4. For FA, after a repeated measures analysis of variance (ANOVA), we did not identify significant interaction between bilateral tracts × groups (Pillai's Trace = 0.011, $F_{[5,127]}$ = 0.295, p=0.915, partial $\eta^2$ = 0.011; Observed Power = 0.122).

| **FA - Tract** | | | **Mean** | **Std. Error** | **95% Confidence Interval** | |
|---|---|---|---|---|---|---|
| | | | | | **Low. Bound** | **Upp. Bound** |
| **Controls** | **Left** | Cingulum | 0.517 | 0.003 | 0.510 | 0.523 |
| | | Corticospinal | 0.614 | 0.002 | 0.610 | 0.619 |
| | | IFO | 0.562 | 0.003 | 0.557 | 0.568 |
| | | ILF | 0.458 | 0.002 | 0.454 | 0.462 |
| | | Parahippocampal | 0.447 | 0.003 | 0.442 | 0.452 |
| | | Uncinate | 0.489 | 0.003 | 0.483 | 0.495 |
| | **Right** | Cingulum | 0.514 | 0.003 | 0.508 | 0.520 |
| | | Corticospinal | 0.613 | 0.002 | 0.609 | 0.618 |
| | | IFO | 0.564 | 0.003 | 0.559 | 0.570 |

| | | | | | | |
|---|---|---|---|---|---|---|
| | | ILF | 0.450 | 0.002 | 0.446 | 0.454 |
| | | Parahippocampal | 0.445 | 0.003 | 0.439 | 0.451 |
| | | Uncinate | 0.488 | 0.003 | 0.482 | 0.494 |
| **Patients** | **Left** | Cingulum | 0.520 | 0.003 | 0.514 | 0.525 |
| | | Corticospinal | 0.616 | 0.002 | 0.613 | 0.620 |
| | | IFO | 0.563 | 0.002 | 0.558 | 0.567 |
| | | ILF | 0.457 | 0.002 | 0.454 | 0.461 |
| | | Parahippocampal | 0.446 | 0.002 | 0.441 | 0.450 |
| | | Uncinate | 0.490 | 0.003 | 0.485 | 0.495 |
| | **Right** | Cingulum | 0.519 | 0.003 | 0.514 | 0.524 |
| | | Corticospinal | 0.613 | 0.002 | 0.609 | 0.616 |
| | | IFO | 0.564 | 0.002 | 0.559 | 0.568 |
| | | ILF | 0.450 | 0.002 | 0.447 | 0.453 |
| | | Parahippocampal | 0.444 | 0.002 | 0.440 | 0.449 |
| | | Uncinate | 0.487 | 0.003 | 0.482 | 0.493 |

*Comparison (Repeated Measures ANOVA) of FA in bilateral tracts of post-COVID patients and controls. Besides no statistical difference, post-COVID patients present a trend of higher FA values in bilateral tracts.*

*FA: Fractional Anisotropy; IFO: Inferior Fronto-Occipital Fasciculus; ILF: Inferior Longitudinal Fasciculus*

**Mean Diffusivity (MD)**

Supplementary Table 5. For MD, after a MANOVA, we did not identify significant interaction between tracts × groups (Pillai's Trace = 0.007, $F_{[4,128]} = 0.237$, p=0.917, partial $\eta^2 = 0.007$, Observed Power = 0.100).

| **MD - Tract** | | **Mean** | **Std. Error** | **95% Confidence Interval** | |
|---|---|---|---|---|---|
| | | | | **Low. Bound** | **Upp. Bound** |
| **Body CC** | **Controls** | 0.0005095 | 0.0000041 | 0.0005013 | 0.0005176 |
| | **Patients** | 0.0005078 | 0.0000034 | 0.0005011 | 0.0005146 |
| **Fornix** | **Controls** | 0.0010973 | 0.0000136 | 0.0010704 | 0.0011243 |
| | **Patients** | 0.0010994 | 0.0000113 | 0.0010771 | 0.0011217 |
| **Genu CC** | **Controls** | 0.0005250 | 0.0000045 | 0.0005162 | 0.0005338 |
| | **Patients** | 0.0005205 | 0.0000037 | 0.0005132 | 0.0005278 |
| **Splenium CC** | **Controls** | 0.0005393 | 0.0000049 | 0.0005296 | 0.0005490 |
| | **Patients** | 0.0005383 | 0.0000041 | 0.0005303 | 0.0005464 |

*Comparison (MANOVA) of MD in midline tracts of post-COVID patients and controls. There is no statistical difference of MD between post-COVID patients and controls.*

*MD: Mean Diffusivity; CC: Corpus Callosum*

Supplementary Table 6. For MD, after a repeated measures ANOVA, we did not identify significant interaction between bilateral tracts × groups (Pillai's Trace = 0.020, $F_{[5,127]}$ = 0.520, p=0.760, partial $\eta^2$ = 0.020; Observed Power = 0.188).

| **MD - Tract** | | | **Mean** | **Std. Error** | **95% Confidence Interval** | |
|---|---|---|---|---|---|---|
| | | | | | **Low. Bound** | **Upp. Bound** |
| **Controls** | **Left** | Cingulum | 0.0007288 | 0.0000035 | 0.0007218 | 0.0007357 |
| | | Corticospinal | 0.0008247 | 0.0000038 | 0.0008172 | 0.0008323 |
| | | IFO | 0.0007970 | 0.0000047 | 0.0007877 | 0.0008063 |
| | | ILF | 0.0007597 | 0.0000032 | 0.0007535 | 0.0007660 |
| | | Parahippocampal | 0.0008309 | 0.0000062 | 0.0008186 | 0.0008432 |
| | | Uncinate | 0.0007776 | 0.0000042 | 0.0007693 | 0.0007859 |
| | **Right** | Cingulum | 0.0007315 | 0.0000036 | 0.0007244 | 0.0007386 |
| | | Corticospinal | 0.0008265 | 0.0000038 | 0.0008191 | 0.0008340 |
| | | IFO | 0.0007986 | 0.0000047 | 0.0007892 | 0.0008079 |
| | | ILF | 0.0007564 | 0.0000032 | 0.0007501 | 0.0007628 |
| | | Parahippocampal | 0.0008315 | 0.0000066 | 0.0008185 | 0.0008445 |
| | | Uncinate | 0.0007783 | 0.0000045 | 0.0007694 | 0.0007871 |
| **Patients** | **Left** | Cingulum | 0.0007238 | 0.0000029 | 0.0007180 | 0.0007295 |
| | | Corticospinal | 0.0008191 | 0.0000032 | 0.0008128 | 0.0008253 |
| | | IFO | 0.0007962 | 0.0000039 | 0.0007885 | 0.0008039 |
| | | ILF | 0.0007542 | 0.0000026 | 0.0007490 | 0.0007594 |
| | | Parahippocampal | 0.0008366 | 0.0000051 | 0.0008265 | 0.0008468 |
| | | Uncinate | 0.0007765 | 0.0000035 | 0.0007696 | 0.0007834 |
| | **Right** | Cingulum | 0.0007232 | 0.0000030 | 0.0007173 | 0.0007290 |
| | | Corticospinal | 0.0008266 | 0.0000031 | 0.0008205 | 0.0008328 |
| | | IFO | 0.0007945 | 0.0000039 | 0.0007868 | 0.0008023 |
| | | ILF | 0.0007525 | 0.0000027 | 0.0007472 | 0.0007578 |
| | | Parahippocampal | 0.0008383 | 0.0000054 | 0.0008276 | 0.0008491 |
| | | Uncinate | 0.0007785 | 0.0000037 | 0.0007712 | 0.0007859 |

*Comparison (Repeated Measures ANOVA) of MD in bilateral tracts of post-COVID patients and controls. There is no statistical difference of MD between post-COVID patients and controls.*

*MD: Mean Diffusivity; IFO: Inferior Fronto-Occipital Fasciculus; ILF: Inferior Longitudinal Fasciculus*

**Radial Diffusivity (RD)**

Supplementary Table 7. For RD, after a MANOVA, we did not identify significant interaction between tracts × groups (Pillai's Trace = 0.006, $F_{[4,128]} = 0.182$, p=0.947, partial $\eta^2 = 0.006$, Observed Power = 0.088).

| **RD - Tract** | | **Mean** | **Std. Error** | **95% Confidence Interval** | |
|---|---|---|---|---|---|
| | | | | **Low. Bound** | **Upp. Bound** |
| **Body CC** | **Controls** | 0.0005095 | 0.0000041 | 0.0005013 | 0.0005176 |
| | **Patients** | 0.0005078 | 0.0000034 | 0.0005011 | 0.0005146 |
| **Fornix** | **Controls** | 0.0010973 | 0.0000136 | 0.0010704 | 0.0011243 |
| | **Patients** | 0.0010994 | 0.0000113 | 0.0010771 | 0.0011217 |
| **Genu CC** | **Controls** | 0.0005250 | 0.0000045 | 0.0005162 | 0.0005338 |
| | **Patients** | 0.0005205 | 0.0000037 | 0.0005132 | 0.0005278 |
| **Splenium CC** | **Controls** | 0.0005393 | 0.0000049 | 0.0005296 | 0.0005490 |
| | **Patients** | 0.0005383 | 0.0000041 | 0.0005303 | 0.0005464 |

*Comparison (MANOVA) of RD in midline tracts of post-COVID patients and controls. Besides no statistical difference, post-COVID patients present a trend of lower RD values in midline tracts.*
*RD: Radial Diffusivity; CC: Corpus Callosum*

Supplementary Table 8. For RD, after a repeated measures ANOVA, we did not identify significant interaction between tracts × groups (Pillai's Trace = 0.022, $F_{[5,127]} = 0.580$, p=0.715, partial $\eta^2 = 0.022$, Observed Power = 0.207).

| **RD - Tract** | | | **Mean** | **Std. Error** | **95% Confidence Interval** | |
|---|---|---|---|---|---|---|
| | | | | | **Low. Bound** | **Upp. Bound** |
| **Controls** | **Left** | Cingulum | 0.0005013 | 0.0000037 | 0.0004941 | 0.0005086 |
| | | Corticospinal | 0.0005110 | 0.0000039 | 0.0005032 | 0.0005187 |
| | | IFO | 0.0005194 | 0.0000040 | 0.0005115 | 0.0005273 |
| | | ILF | 0.0005605 | 0.0000030 | 0.0005546 | 0.0005664 |
| | | Parahippocampal | 0.0006206 | 0.0000055 | 0.0006098 | 0.0006314 |
| | | Uncinate | 0.0005557 | 0.0000045 | 0.0005468 | 0.0005646 |
| | **Right** | Cingulum | 0.0005052 | 0.0000037 | 0.0004978 | 0.0005126 |
| | | Corticospinal | 0.0005130 | 0.0000039 | 0.0005053 | 0.0005206 |
| | | IFO | 0.0005193 | 0.0000041 | 0.0005111 | 0.0005275 |
| | | ILF | 0.0005631 | 0.0000030 | 0.0005572 | 0.0005690 |
| | | Parahippocampal | 0.0006220 | 0.0000060 | 0.0006103 | 0.0006338 |
| | | Uncinate | 0.0005568 | 0.0000048 | 0.0005474 | 0.0005662 |
| | | Cingulum | 0.0004966 | 0.0000030 | 0.0004906 | 0.0005026 |

| | | | | | | |
|---|---|---|---|---|---|---|
| **Patients** | **Left** | Corticospinal | 0.0005056 | 0.0000032 | 0.0004992 | 0.0005120 |
| | | IFO | 0.0005194 | 0.0000033 | 0.0005129 | 0.0005259 |
| | | ILF | 0.0005575 | 0.0000025 | 0.0005526 | 0.0005624 |
| | | Parahippocampal | 0.0006261 | 0.0000045 | 0.0006171 | 0.0006350 |
| | | Uncinate | 0.0005549 | 0.0000037 | 0.0005475 | 0.0005623 |
| | **Right** | Cingulum | 0.0004967 | 0.0000031 | 0.0004906 | 0.0005028 |
| | | Corticospinal | 0.0005134 | 0.0000032 | 0.0005070 | 0.0005197 |
| | | IFO | 0.0005179 | 0.0000034 | 0.0005111 | 0.0005247 |
| | | ILF | 0.0005606 | 0.0000025 | 0.0005557 | 0.0005655 |
| | | Parahippocampal | 0.0006282 | 0.0000049 | 0.0006184 | 0.0006379 |
| | | Uncinate | 0.0005579 | 0.0000039 | 0.0005501 | 0.0005657 |

*Comparison (Repeated Measures ANOVA) of RD in bilateral tracts of post-COVID patients and controls. Besides no statistical difference, post-COVID patients present a trend of lower RD values in bilateral tracts.*
*RD: Radial Diffusivity; IFO: Inferior Fronto-Occipital Fasciculus; ILF: Inferior Longitudinal Fasciculus*

**Axial Diffusivity (AD)**

Supplementary Table 9. For AD, after a MANOVA, we did not identify significant interaction between tracts × groups (Pillai's Trace = 0.010, $F_{[4,128]} = 0.334$, p=0.855, partial $\eta^2 = 0.010$, Observed Power = 0.124).

| **AD - Tract** | | **Mean** | **Std. Error** | **95% Confidence Interval** | |
|---|---|---|---|---|---|
| | | | | **Low. Bound** | **Upp. Bound** |
| **Body CC** | **Controls** | 0.0013508 | 0.0000079 | 0.0013352 | 0.0013664 |
| | **Patients** | 0.0013582 | 0.0000065 | 0.0013453 | 0.0013711 |
| **Fornix** | **Controls** | 0.0021418 | 0.0000191 | 0.0021040 | 0.0021797 |
| | **Patients** | 0.0021501 | 0.0000158 | 0.0021188 | 0.0021814 |
| **Genu CC** | **Controls** | 0.0012870 | 0.0000064 | 0.0012743 | 0.0012997 |
| | **Patients** | 0.0012855 | 0.0000053 | 0.0012750 | 0.0012960 |
| **Splenium CC** | **Controls** | 0.0014630 | 0.0000085 | 0.0014462 | 0.0014798 |
| | **Patients** | 0.0014696 | 0.0000070 | 0.0014557 | 0.0014835 |

*Comparison (MANOVA) of AD in midline tracts of post-COVID patients and controls. Besides no statistical difference, post-COVID patients present a trend of lower AD values in midline tracts.*
*AD: Axial Diffusivity; CC: Corpus Callosum*

Supplementary Table 10. For AD, after a repeated measures ANOVA, we did not identify significant interaction between tracts × groups (Pillai's Trace = 0.020, $F_{[5,127]}$ = 0.517, p=0.763, partial $\eta^2$ = 0.020, Observed Power = 0.187).

| **AD - Tract** | | | **Mean** | **Std. Error** | **95% Confidence Interval** | |
|---|---|---|---|---|---|---|
| | | | | | **Low. Bound** | **Upp. Bound** |
| **Controls** | **Left** | Cingulum | 0.0011835 | 0.0000059 | 0.0011718 | 0.0011952 |
| | | Corticospinal | 0.0014523 | 0.0000048 | 0.0014427 | 0.0014619 |
| | | IFO | 0.0013522 | 0.0000079 | 0.0013366 | 0.0013679 |
| | | ILF | 0.0011581 | 0.0000047 | 0.0011489 | 0.0011674 |
| | | Parahippocampal | 0.0012517 | 0.0000085 | 0.0012350 | 0.0012684 |
| | | Uncinate | 0.0012214 | 0.0000050 | 0.0012115 | 0.0012312 |
| | **Right** | Cingulum | 0.0011841 | 0.0000058 | 0.0011727 | 0.0011955 |
| | | Corticospinal | 0.0014539 | 0.0000048 | 0.0014444 | 0.0014635 |
| | | IFO | 0.0013570 | 0.0000076 | 0.0013419 | 0.0013721 |
| | | ILF | 0.0011431 | 0.0000047 | 0.0011339 | 0.0011523 |
| | | Parahippocampal | 0.0012506 | 0.0000087 | 0.0012334 | 0.0012678 |
| | | Uncinate | 0.0012213 | 0.0000051 | 0.0012112 | 0.0012314 |
| **Patients** | **Left** | Cingulum | 0.0011783 | 0.0000049 | 0.0011687 | 0.0011880 |
| | | Corticospinal | 0.0014460 | 0.0000040 | 0.0014381 | 0.0014539 |
| | | IFO | 0.0013495 | 0.0000066 | 0.0013366 | 0.0013625 |
| | | ILF | 0.0011476 | 0.0000039 | 0.0011400 | 0.0011553 |
| | | Parahippocampal | 0.0012576 | 0.0000070 | 0.0012438 | 0.0012714 |
| | | Uncinate | 0.0012198 | 0.0000041 | 0.0012117 | 0.0012279 |
| | **Right** | Cingulum | 0.0011758 | 0.0000048 | 0.0011664 | 0.0011853 |
| | | Corticospinal | 0.0014531 | 0.0000040 | 0.0014452 | 0.0014610 |
| | | IFO | 0.0013479 | 0.0000063 | 0.0013354 | 0.0013604 |
| | | ILF | 0.0011363 | 0.0000038 | 0.0011287 | 0.0011439 |
| | | Parahippocampal | 0.0012586 | 0.0000072 | 0.0012444 | 0.0012728 |
| | | Uncinate | 0.0012197 | 0.0000042 | 0.0012114 | 0.0012281 |

*Comparison (Repeated Measures ANOVA) of AD in midline tracts of post-COVID patients and controls. Besides no statistical difference, post-COVID patients present a trend of lower AD values in bilateral tracts.*
*AD: Axial Diffusivity; IFO: Inferior Fronto-Occipital Fasciculus; ILF: Inferior Longitudinal Fasciculus*

### FUNCTIONAL CONNECTIVITY RESULTS

#### Supplementary Table 11. T-test of patients and controls

| cluster | cluster | Peak | | | |
|---|---|---|---|---|---|
| p(FDR-corr) | Cluster Size | T | p(unc) | x,y,z {mm} | Region of Interest |

| | | | | | | | |
|---|---|---|---|---|---|---|---|
| 0.084 | 31 | 4.535 | <0.001 | 30 | -70 | 26 | Right Superior/Middle Occipital Gyrus |
| 0.063 | 40 | 4.264 | <0.001 | -44 | 4 | 2 | Left Anterior Insula |
| *T-test of Posterior Cingulate Cortex (a Default Mode Network hub) functional connectivity map of post-COVID patients and controls. Observe the trend forward a higher connectivity of Posterior Cingulate Cortex and Left Insula and Right Occipital cortex.* | | | | | | | |

**Supplementary Table 12. Results from the regression between maps of DMN and scores of Fatigue.**

| Cluster | | Peak | | | | | |
|---|---|---|---|---|---|---|---|
| p(FDR-corr) | Cluster Size | T | p(unc) | x,y,z {mm} | | | Region of Interest |
| 0.016 | 199 | 4.582 | <0.001 | -46 | -64 | 36 | Left Angular Gyrus |
| | | 3.897 | <0.001 | -44 | -58 | 26 | Left Angular Gyrus |
| 0.006 | 57 | 4.294 | <0.001 | 42 | -56 | 22 | Right Angular Gyrus |
| *Linear regression of Posterior Cingulate Cortex (a Default Mode Network hub) functional connectivity with fatigue in post-COVID patients. Higher levels of fatigue correlate with a higher connectivity of Posterior Cingulate Cortex with Bilateral Angular Gyri.*<br>*FDRc=57* | | | | | | | |

**Supplementary Table 13.Results from the regression between maps of DMN and scores of excessive Sleepiness.**

| Cluster | | Peak | | | | | |
|---|---|---|---|---|---|---|---|
| p(FDR-corr) | Cluster Size | T | p(unc) | x,y,z {mm} | | | Region of Interest |
| 0.016 | 54 | 5.052 | <0.001 | -8 | -20 | 18 | Left Thalamus |
| | | 3.614 | <0.001 | -8 | -28 | 16 | Left Thalamus |
| 0.025 | 42 | 4.753 | <0.001 | -22 | 10 | 0 | Left Putamen |
| *Linear regression of Posterior Cingulate Cortex (a Default Mode Network hub) functional connectivity with sleepiness in post-COVID patients. Higher levels of sleepiness correlate with a higher connectivity of Posterior Cingulate Cortex with Left Thalamus and Left Putamen..*<br>*FDRc=42* | | | | | | | |

**Supplementary Table 14. Online questionnaire - SECTION 1 / 2**

We are researchers from the Department of Neurology (School of Medical Sciences / UNICAMP) and from the Department of Biology (UNICAMP), and we are studying the effects of coronavirus in the central nervous system.

This questionnaire will help us to understand how people are recovering themselves after the infection by the new coronavirus. Our complete project includes a magnetic resonance, neurological and cognitive examination (memory, language…). If it is possible for you to answer this questionnaire, we would be very grateful.

Principal investigator: Prof. Clarissa Lin Yasuda (CRM 94104)

Contact: (19) 99768-7517

*Required

| | | | | |
|---|---|---|---|---|
| **E-mail address*** | (Your e-mail) | | | |
| **Name*** | (Your answer) | | | |
| **Age*** | (Your Answer) | | | |
| **Gender** | Male | Female | I prefer not to answer | Other: |
| **Phone** | (Your answer) | | | |
| **City / State** | (Your answer) | | | |

| | | | | |
|---|---|---|---|---|
| **Do you have any association with UNICAMP?**<br><br>(Are you a student, professor or worker at UNICAMP?) | Yes | No | | |
| **Date of diagnosis** (COVID infection)* | Date of confirmation (with a test) | | | |
| **Diagnostic Method**<br><br>What was the confirmation's method of the Covid diagnosis? | PCR (Swab) | Antibodies (Blood test / Quick test) | Antibody + PCR | Other: |
| **Treatment**<br><br>Select the type of treatment for the COVID infection | Home | Hospital - Infirmary | Hospital – Intensive Care Unit | |
| **Symptoms of acute infection***<br><br>Describe the symptoms you presented during the acute period (in the hospital or individual isolation, in case you had the treatment done). Select all the relevant symptoms. | Shortness of breath | Tiredness / Fatigue | Fever | |
| | Olfactory changes | Headache | Taste changes | |
| | No symptoms | Other: ______________________________ | | |

| **Symptoms in the first month after hospital discharge / quarantine**<br><br>Describe the symptoms in the period after the hospital discharge or after the end of individual isolation (If you had your treatment at home). Select all the relevant symptoms. | Shortness of breath | Tiredness / Fatigue | Fever |
|---|---|---|---|
| | Olfactory changes | Headache | Taste changes |
| | No symptoms | Other: | |

After section 1, continue to section 2

**SECTION 2 / 2**

| **SYMPTOMS AFTER COVID INFECTION**<br><br>In this section, we will talk about the symptoms that occurred after the recovering of COVID 19 infection. | | | |
|---|---|---|---|
| **Frequent Post-Covid Symptoms**<br><br>Check the symptoms that had after the | Headache | Olfactory changes | Taste changes |
| | | Shortness of breath | Fever |

| | | | |
|---|---|---|---|
| infection, but may not necessarily persisted up to now. | Tiredness / Fatigue | | |
| | Somnolence during the day | Memory problems | Difficulties with daily activities |
| | Motor Difficulties | Coordination difficulties | I do not present any symptom |
| | Other:___________________________ | | |
| **Current Post-Covid Symptoms** Check the currently persistent symptoms you had after the infection | Headache | Olfactory changes | Taste changes |
| | Tiredness / Fatigue | Shortness of breath | Fever |
| | Somnolence during the day | Memory problems | Difficulties with daily activities |

| | | | |
|---|---|---|---|
| | Motor Difficulties | Coordination difficulties | I do not present any symptom |
| | Other:________________________________ | | |
| **Would you like to describe any more symptoms?** | (Your answer) | | |
| **May we contact you?** | Yes | No | |
| **If yes, do you have a preferred medium of contact?** | Registered Email | Registered Phone | Other |
